# A Cohort Study of Urinary Tract Infections in People Living with Dementia: Epidemiology and Diagnostic Challenges

**DOI:** 10.1101/2024.12.10.24318775

**Authors:** Raphaella Jackson, Rory Cave, Martin Tran, Kirsten Jensen, Michael A. Crone, Alexander J. Webb, Loren P. Cameron, Ramin Nilforooshan, David Wingfield, David J. Sharp, Paul S. Freemont

## Abstract

**Background:** Urinary tract infections (UTIs) are a leading cause of hospitalisation in people living with dementia (PLWD), making accurate detection and prompt treatment essential in this vulnerable population.

**Methods:** This retrospective longitudinal cohort study assessed the concordance between self-reported symptoms, urine colony counts >10□ CFU/mL, dipstick results positive for leukocytes and/or nitrites, and urinary IL-8 levels in identifying UTIs among PLWD. The study included 78 community-dwelling individuals aged over 50 with a confirmed dementia diagnosis, recruited from cohorts established by the Surrey and Borders Partnership NHS Foundation Trust and the Hammersmith & Fulham Partnership Primary Care Network between late 2019 and 2023.

**Results:** UTI frequency among PLWD was highly variable, with some individuals experiencing recurrent infections whilst others had none throughout the study period. The microbial taxa identified were consistent with those seen in other populations. There was no clear concordance between self-reported symptoms and laboratory indicators of UTI. However, dipstick-positive results correlated with urine samples showing >10□ CFU/mL of a single colony morphology growth and elevated IL-8 concentrations.

**Conclusions:** Urinary dipstick tests for nitrites and leukocytes may serve as a practical screening tool for UTIs in PLWD, particularly in individuals unable to reliably report symptoms. However, future research is needed to evaluate the clinical impact of this diagnostic approach on outcomes such as hospitalisation rates, delirium incidence, and antibiotic resistance and stewardship in this vulnerable population.

## Background

At any given time, one in four hospital beds in the United Kingdom is occupied by people living with dementia (PLWD). Around 20% of these admissions result from acute conditions, such as infections, where timely intervention could prevent serious outcomes [1, 2]. Among these, urinary tract infections (UTIs) are especially common, accounting for approximately 9% of hospital admissions in this population [2–4]. Beyond hospitalisation risk, delayed UTI diagnosis in PLWD can lead to increased confusion, delirium, sepsis, faster clinical deterioration [5–7], and potentially worsened neurodegeneration [8, 9].

A major barrier to timely UTI detection in PLWD is symptom presentation. Current UK and European guidelines recommend diagnosing and treating UTIs only when localised signs or symptoms are evident [10, 11]. However, infection symptoms are often atypical or absent in older adults, particularly for UTIs [12, 13] and PLWD may struggle to recognise or communicate symptoms due to cognitive impairment [14]. Reliable biomarkers for diagnosing UTIs in the absence of symptoms are currently lacking. Moreover, the presence of bacteria in urine (asymptomatic bacteriuria) is common among the elderly [15], and treating it has not shown clinical benefit [16–18].

While UTIs have been studied extensively across various populations, little is known about which uropathogens affect PLWD specifically. Given both the impact on individual well-being and the broader public health burden, there is a clear need to better understand UTIs in this group. In this study, we conducted a longitudinal analysis of urinary pathogens in people with Alzheimer’s disease, the most prevalent form of dementia, who were still living at home. By combining molecular, microbiological, and clinical data, we aimed to characterise bacterial colonisation and infection in this understudied, community-dwelling population, filling a gap in a field that has focused largely on institutionalised cohorts.

## Methods

### Study Cohort

The cohort for this study was recruited between late 2019 and December 2023 through the Surrey and Borders Partnership NHS Foundation Trust (SABP) and the Hammersmith & Fulham Partnership Primary Care Network (HFP PCN). Individuals with a confirmed diagnosis of dementia (regardless of subtype) were enlisted via NHS Community Mental Health Teams for Older People (CMHT-OP), SABP’s specialist memory services, and equivalent services in North West London. Recruitment was further supported by promotion on the national Join Dementia Research platform (https://www.joindementiaresearch.nihr.ac.uk/).

#### Inclusion Criteria

- Aged 50 or older at baseline
- Confirmed dementia diagnosis by specialist assessment
- Able and willing to provide informed consent
- Standardised Mini-Mental State Examination (SMMSE) score at baseline interview over 12
- Living in the community
- Sufficient functional English to complete assessments

#### Exclusion Criteria

- Living in a residential care home
- Unstable mental state (e.g., severe depression, psychosis, agitation, anxiety, or suicidal ideation)
- Severe sensory impairment
- SMMSE score <12
- Unable to communicate verbally
- Under treatment for a terminal illness

### Urine Collection

Urine samples were collected every 4–6 weeks using sterile containers or, for those with fine motor difficulties, via a sterile kidney dish. Samples were transported on ice for analysis.

### Chromogenic Agar Plates

To quantify bacterial colonies, 1 μL of urine mixed with 99 μL 1× sterile PBS was cultured on Brilliance UTI Clarity Agar (Oxoid Ltd) and incubated at 37°C for 18–24 hours. Colony counts were recorded as CFU/mL, capped at 100,000+ CFU/mL.

### Overnight Cultures of Bacterial Strains

Selected colonies were cultured overnight in Tryptone Soy Broth (Oxoid Ltd) at 37°C with shaking at 200 rpm. Subsequently, a 400 μL aliquot of culture was mixed with 400 μL of sterile 50% glycerol in 2D barcoded matrix tubes (Nunc™) and stored at -80°C.

### 16S rRNA Gene Amplification, Sequencing, and Classification

To confirm bacterial identity, 1 μL of overnight culture was mixed with 99 μL 1× sterile PBS and heated at 95°C for 30 minutes, after which the V3-V4 regions of the 16S rRNA gene were amplified using:

- **Forward primer:** AGGGTTTTCCCAGTCACGACGTTCCTACGGGNGGCWGCA
- **Reverse primer:** GACTACHVGGGTATCTAATCC

PCR conditions: 1 cycle of 95°C for 5 min; 30 cycles of 95°C for 30 s, 50°C for 30 s, 1 cycle of 72°C for 30 s; final extension at 72°C for 5 min.

Amplicons (5 μL) were treated with 2 μL ExoSAP-IT™ at 37°C for 4 min, then 80°C for 1 min. Products were sent to Eurofins Genomics for Sanger sequencing using the M13 uni (-43) primer (AGGGTTTTCCCAGTCACGACGTT). Sequences were quality-filtered (QV30) and classified via the SILVA ACT tool using default settings.

### Interleukin 8 (IL-8) Concentration in Urine

Urine samples were centrifuged at >500 × *g* for 10 minutes at 4⍰°C to remove insoluble material. Supernatants were diluted 1:4 with Olink® Focus Sample Diluent to a final volume of 50⍰μL and stored at -80⍰°C until shipment. Samples were submitted to the UKDRI Fluid Biomarker Laboratory and Biomarker Factory for cytokine quantification using the Olink® Target 48 inflammation panel on the Olink® Signature Q100 instrument. This panel employs Olink’s Proximity Extension Assay (PEA) technology, in which matched antibody pairs conjugated to DNA oligonucleotides bind to the target protein, allowing hybridisation and extension by polymerase. This enables highly sensitive and specific protein quantification via qPCR.

### Data Pre-Processing and Classification

#### Symptom Reporting

Participants completed a UTI symptom questionnaire (Supplementary Table 1) on the day of sample collection. Their symptom responses were manually reviewed and recoded as follows:

1. Responses with a clear yes/no intent, including synonyms or elaborations, were coded accordingly.
2. Non-responses, unrelated causes, or expressions of uncertainty were coded as “Other”.

Supplementary Table 1 provides details of the original and recoded responses.

#### UTI Classification

UTI was defined as the presence of bacterial growth with a single colony morphology exceeding 10□ CFU/mL, accompanied by the detection of leukocytes and/or nitrites [19, 20]. Symptomatic status was based on an adapted questionnaire per NICE guidelines [19].

### Data Analysis

All analyses were conducted in R. Descriptive statistics summarised cohort demographics and UTI frequency. Associations between laboratory indicators, bacterial taxa, symptoms, and UTI frequency were assessed using generalized linear mixed-effects models (GLMMs) with a logit link, with separate models applied for distinct analyses. Repeated measures were included as random effects. Odds ratios (ORs) with 95% confidence intervals (CIs) were reported. False discovery rate (FDR) correction adjusted for multiple comparisons models. Bias-reduced binomial GLMMs explored associations with participants with High Frequency of UTI. Linear mixed-effects models (LMMs) quantified relationships between IL-8 levels, bacterial growth, dipstick results, and symptoms. Sensitivity and specificity calculations were performed at both the participant and sample levels to evaluate the diagnostic performance of symptom reporting, dipstick testing, and bacterial counts. UTI frequency was categorised per 6-week windows into: “No UTI,” “Low-Frequency,” “Mid-Frequency,” or “High-Frequency”. Analysis code and outputs are available in the supplementary materials.

## Results

### Cohort characteristic

Over the four-year study period, 602 urine samples were collected approximately every six weeks from 86 participants (Figure 1). All participants were living at home, and 55% lived with a carer. The cohort was nearly gender-balanced (45% female), with Alzheimer’s disease being the most common diagnosis (66%). The median age at enrolment was 82 years (interquartile range [IQR]: 77–88). Due to rolling recruitment, participants contributed varying numbers of samples, with a mean of seven samples per person (IQR: 3–10).

**Figure 1:**
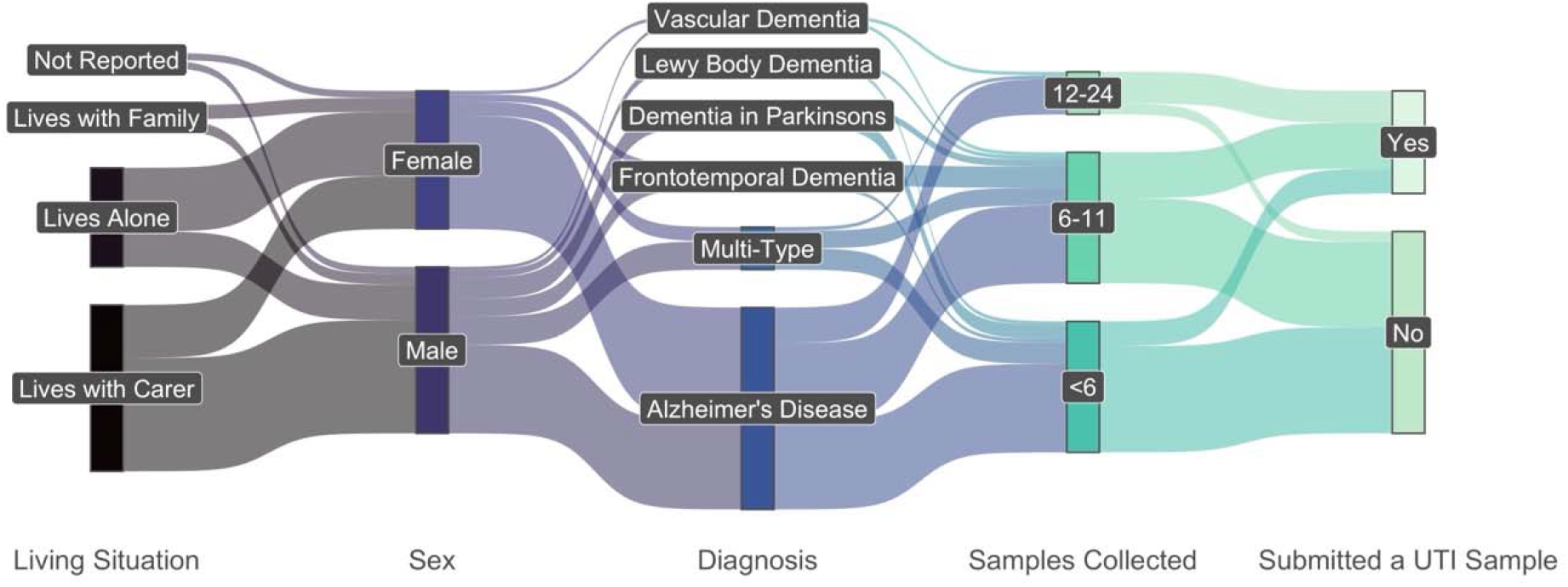
Participant Demographics and Samples Collected. A Sankey plot showing the study cohort’s living situation, gender, diagnosis, number of samples collected per participant, and whether they submitted at least one UTI sample.

Clinicians classified 512 samples as non-UTI and 90 as UTI. Of the UTI-positive samples, 65% were from females and 35% from males. Overall, 33% of participants submitted at least one UTI-positive sample (Figure 1).

### Variable UTI frequency among PLWD

To assess how often participants experienced UTIs, we grouped individuals based on the occurrence of UTI events within six-week intervals—the approximate time between sample collections (Figure 2). A window was marked “positive” if a UTI was recorded during that period. We focused this analysis on 58 participants who were enrolled for at least six months; the remaining 28 participants were classified as “Short-Term” and excluded from frequency-based categorisation.

**Figure 2:**
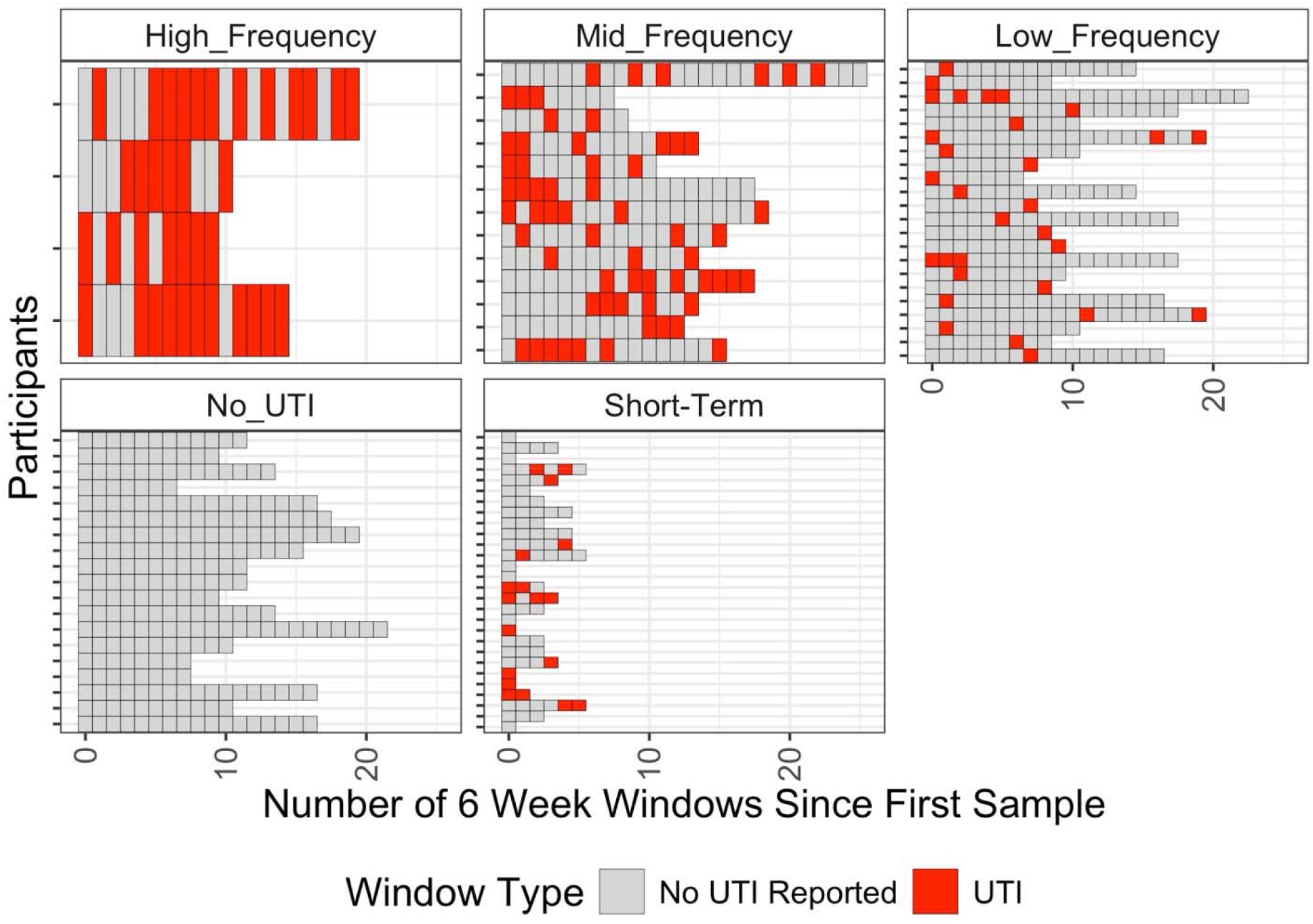
UTI Frequency in Cohort. A plot showing UTI occurrence in each 6-week window following a participant’s first sample. Participants are grouped by UTI Frequency Type. Those with less than 6 months of study participation are labelled “Short-Term” and not further classified. The X-axis represents sequential 6-week intervals since each participant’s first sample, with each unit indicating whether a UTI event occurred within that window. The Y-axis represents individual participants. The figure demonstrates substantial variability in UTI frequency between participants.

Participants with no positive windows were classified as “No UTI”. Those with fewer than 20% of their windows marked positive were labelled “Low-frequency”, those with 20–50% as “mid-frequency”, and those with more than 50% as “high-frequency”. In total, we identified 4 high-frequency, 13 mid-frequency, 22 low-frequency, and 19 no-UTI participants.

### Microbiological Characteristics of Collected Samples

To characterise the chemical and microbial composition of each urine sample, we performed urinary dipstick testing and bacterial culturing on chromogenic agar—both standard clinical methods for detecting UTIs [19, 20]. For the dipstick analysis, we focused exclusively on the presence of leukocytes and nitrites, excluding other markers from consideration (Figure 3A)[21]. Of the 602 samples collected, 597 underwent dipstick testing. Among these, 26% were positive for leukocytes and 8% for nitrites. As not all bacteria produce nitrites, their absence does not exclude infection.

**Figure 3:**
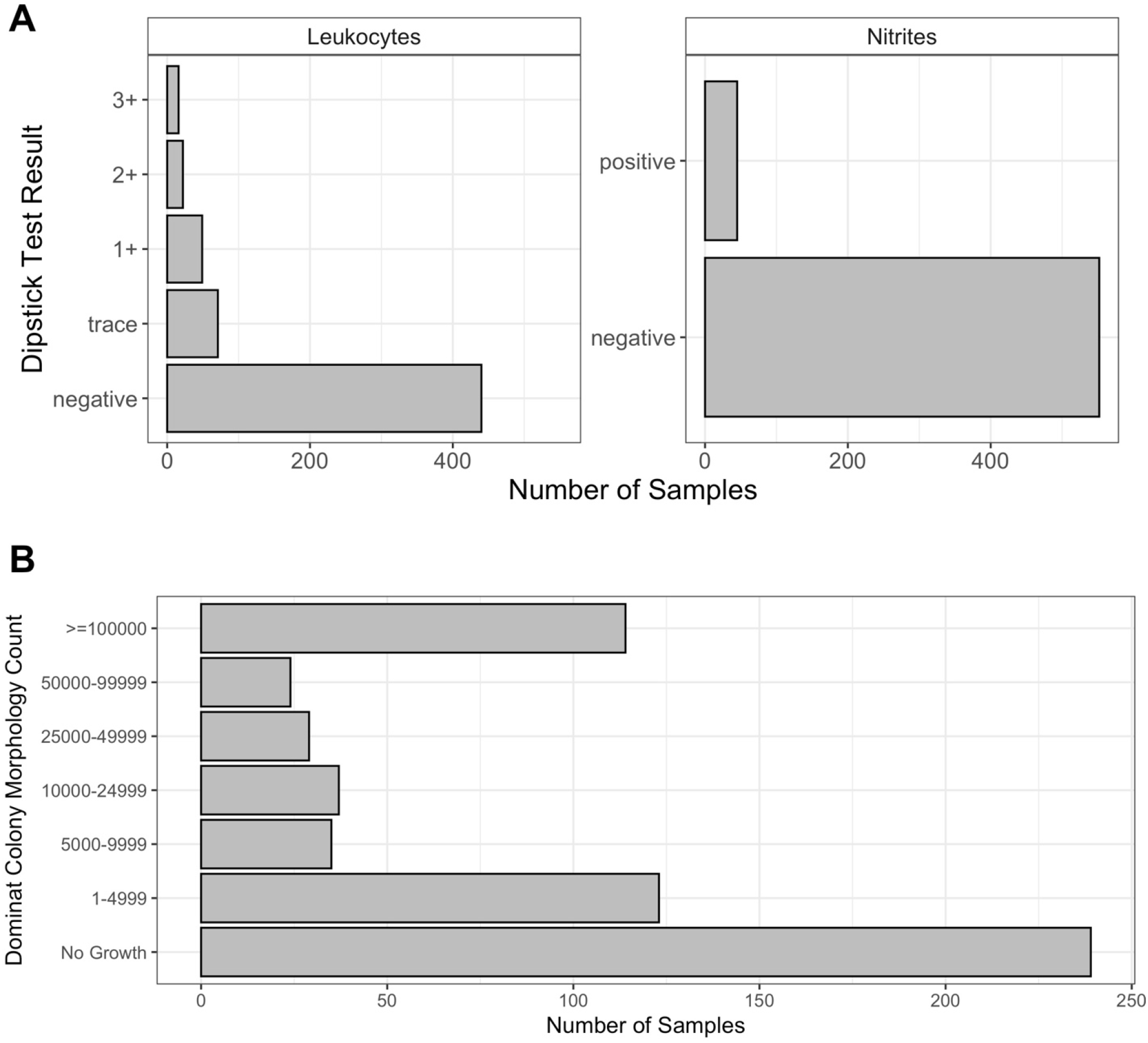
Dipstick Test Results and Dominant Colony Morphology Count. (A) Bar plot displaying leukocyte and nitrite results from dipstick tests across all analysable urine samples. (B) Bar plot showing the colony-forming unit (CFU/mL) count of the most dominant colony morphology observed on chromogenic agar following culture of each sample. The figure indicates that the majority of samples do not exhibit strong evidence of urinary tract infection, as reflected by negative dipstick results for leukocytes and nitrite, alongside culture growth below the diagnostic threshold of 10^5^ CFU/mL.

Bacterial growth was detected in 362 samples, with one excluded due to excessive mixed growth, leaving 361 for colony counting (Figure 3B). Using the UK NICE threshold of ≥10□ CFU/mL for a single colony morphology to define UTIs [19], 19% of samples met this criterion. A generalised linear mixed model (GLMM) revealed a strong association between a positive dipstick result and culture positivity above this threshold OR = 18.1, p < 0.001). The dipstick test sensitivity —defined as a positive dipstick result in the presence of a single colony morphology ≥10□ CFU/mL— was 82% per participant and 70% per sample. Dipstick test specificity— defined as a negative dipstick result in the absence of a single colony morphology ≥10□ CFU/mL—was 73% per participant and 84% per sample. This indicates that while a positive dipstick result substantially increases the likelihood of a culture-confirmed UTI, dipstick tests are slightly more effective at ruling out samples without high bacterial loads than they are at reliably identifying those with high bacterial loads — particularly when assessed at the sample level.

### Common Uropathogens Detected in PLWD and Their Association with Laboratory

#### Markers

To determine whether cultured bacteria from urine samples were known uropathogens, we performed 16S rRNA gene Sanger sequencing on a subset of 330 isolates out of 771 from the 362 samples with bacterial growth. We then compared the taxonomic profiles of isolates from dipstick-positive samples (leukocytes and/or nitrites) that exceeded the ≥10^5^ CFU/mL threshold with those that did not meet these criteria. Given known sex-related differences in urinary tract colonisation, participant sex was included as a variable but was insignificant (p = 0.5) in our analysis (Figure S1) [22, 23].

*Escherichia* was the most frequently identified genus, consistent with previous studies, and was strongly associated with both dipstick positivity and high colony counts (OR = 16.4, p < 0.001, FDR-adjusted p < 0.001) [24]. *Klebsiella* (OR = 75.4, p < 0.001, FDR-adjusted p < 0.001) and *Streptococcus* (OR = 9.6, p = 0.003, FDR-adjusted p = 0.004) also showed strong associations, supporting their roles as common uropathogens [24].

Although *Enterococcus* and *Staphylococcus* were not significantly associated in our model, they were frequently detected in dipstick-negative samples. As members of the gut and skin microbiota, their presence may reflect low-level contamination during sampling, increased susceptibility to colonisation in this cohort, or both. Nonetheless, since these genera can cause UTIs and were observed in participants classified as UTI-positive, their potential clinical relevance should not be overlooked.

To prevent complete separation when modelling dipstick results—in which samples with multiple organisms have the same outcome—we restricted the analysis to the dominant taxon per sample. Klebsiella was excluded from this primary analysis, as all dominant Klebsiella samples tested dipstick-positive. In this context, both Escherichia (OR = 12.06, p < 0.001, FDR-adjusted p < 0.001) and Streptococcus (OR = 9.7, p = 0.008, FDR-adjusted p = 0.014) were significantly associated with dipstick positivity. When Klebsiella and multiple taxa per sample were included and focusing on samples with ≥10^5^⍰CFU/mL of a single colony type, Escherichia (OR = 5.9, p < 0.001, FDR-adjusted p < 0.001) and Klebsiella (OR = 19.0, p < 0.001, FDR-adjusted p < 0.001) remained significant. These results suggest that dipstick tests reliably detect samples dominated by potentially pathogenic taxa, particularly Escherichia and Klebsiella, when present at ≥10^5^⍰CFU/mL.

### No Association Between Bacterial Taxa and UTI Frequency in PLWD

To explore potential microbial drivers of recurrent infection, we examined whether specific bacterial taxa were associated with differing frequencies of presumed UTIs across participants (Figure S2). While we hypothesised that certain species might underlie high - frequency of infections, our *logistic* model found no clear associations—likely due to limited statistical power, as only four participants met the criteria for high-frequency of UTI. Additionally, participants with more frequent infections were not consistently colonised by a single bacterial genus. For example, some individuals (e.g., P061 and P004) were predominantly infected by a single genus across timepoints, whereas others (e.g., P054) exhibited greater variability, with different dominant isolates detected in different samples.

### Symptom Reporting Does Not Predict Laboratory Evidence of UTI

Given that self-reported symptoms form the primary basis for diagnosing UTIs in individuals over 65 according to NICE guidelines, we assessed the relationship between reported symptoms and laboratory indicators of infection (Figure 4) [19].

**Figure 4:**
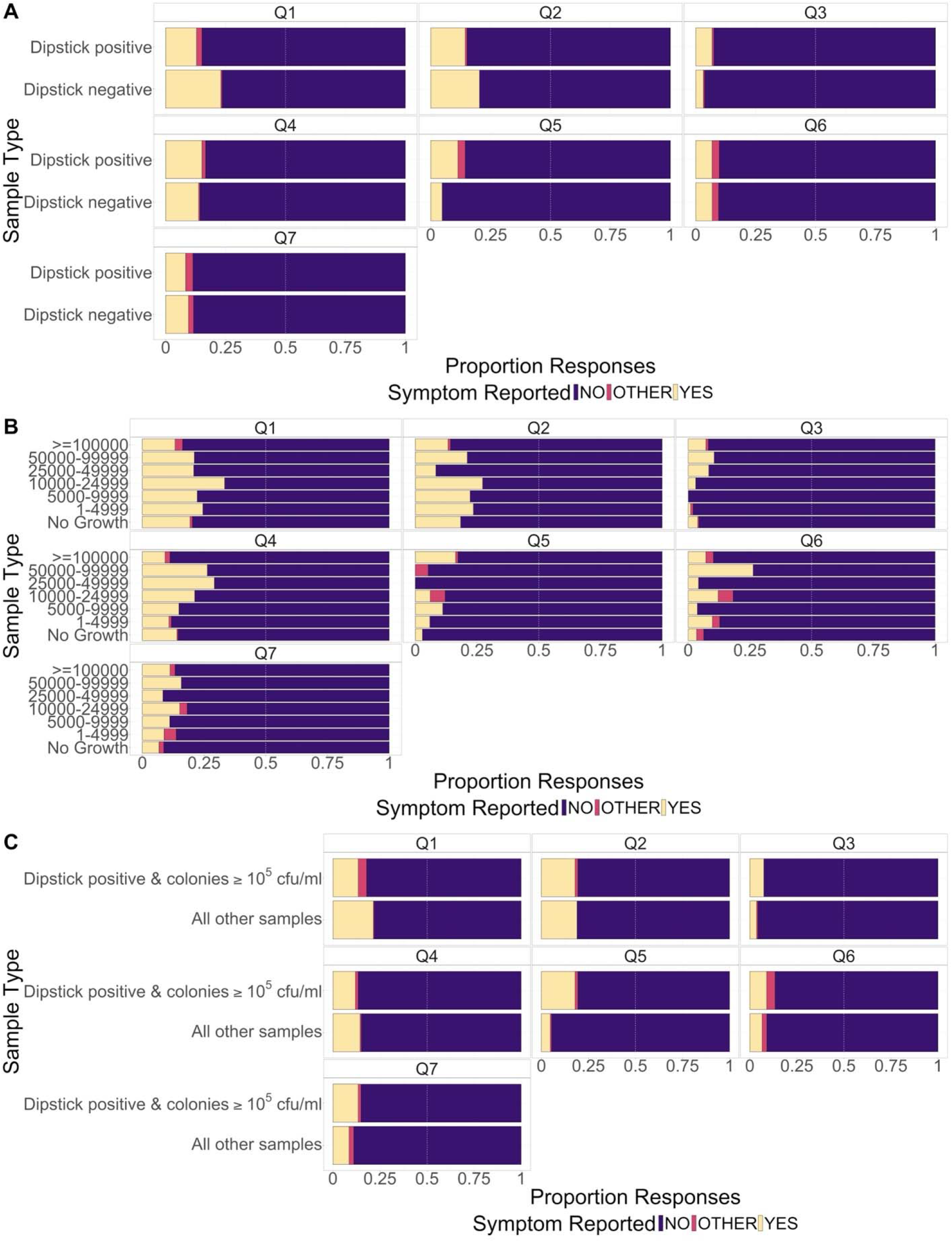
Symptom Reporting by Dipstick Result and Colony Count. (A) Proportion of reported symptoms stratified by dipstick result type. (B) Proportion of reported symptoms stratified by colony count.(C) Proportion of reported symptoms based on the combination of a dipstick-positive result and ≥100,000 CFU/mL culture growth. The figure shows no clear association between symptom questionnaire responses and either dipstick test results or culture findings.

At each visit, participants answered seven questions addressing common UTI symptoms, including increased urgency, frequency, dysuria, urinary retention, changes in urine smell or colour, localised pain, and general malaise. A “yes” response to any question classified the individual as symptomatic.

Most participants reported no symptoms across all categories. There was no significant association between symptom presence and dipstick positivity (p = 0.2). Sensitivity—defined as the proportion of symptomatic participants with a positive laboratory test—was 45% per participant and 29% per sample, while specificity—defined as the proportion of asymptomatic participants with a negative laboratory test—was 46% per participant and 76% per sample. High bacterial counts (>10□ CFU/mL) likewise showed no significant link to symptoms (p = 0.7), with the same sensitivity (45% per participant; 29% per sample) and specificity (46% per participant; 76% per sample). Combining dipstick positivity and high colony counts also failed to reveal a relationship with symptoms (p = 0.4), producing a sensitivity of 30% per participant and 29% per sample, and a specificity of 73% per participant and 76% per sample. These results indicate that self-reported symptoms alone lack the sensitivity required to detect laboratory-confirmed infection, so relying solely on symptom reports would miss a substantial number of cases.

### Dipstick Results Correlate with Inflammatory Marker IL-8

To evaluate the biological relevance of laboratory-defined UTIs, we examined a subset of data from 31 participants (107 samples) using a linear mixed model (LMM). This analysis assessed associations between urinary IL-8—a validated biomarker of inflammation in UTIs[25]—and bacterial colony counts for different taxon. A second, separate LMM of IL-8 on dipstick results and self-reported symptoms was employed to avoid over-adjustment and collider bias with bacterial counts, reduce multicollinearity, isolate the clinical proxy evaluation from the mechanistic model, and maximise statistical power.

IL-8 levels were significantly associated with log-transformed colony counts of *Escherichia* (coefficient = 0.12, p = 0.005, FDR-adjusted p = 0.01), *Klebsiella* (coefficient = 0.19, p = 0.011, FDR-adjusted p = 0.016), *Staphylococcus* (coefficient = 0.15, raw p < 0.001, FDR-adjusted p < 0.001), and *Enterococcus* (coefficient = 0.13, p = 0.015, FDR-adjusted p = 0.018). Dipstick positivity (leukocytes and/or nitrites) was also strongly linked to elevated IL-8 levels (coefficient = 2.2, p < 0.001, FDR-adjusted p < 0.001). In contrast, self-reported symptoms showed no significant association with IL-8 concentrations (Figure 5).

**Figure 5:**
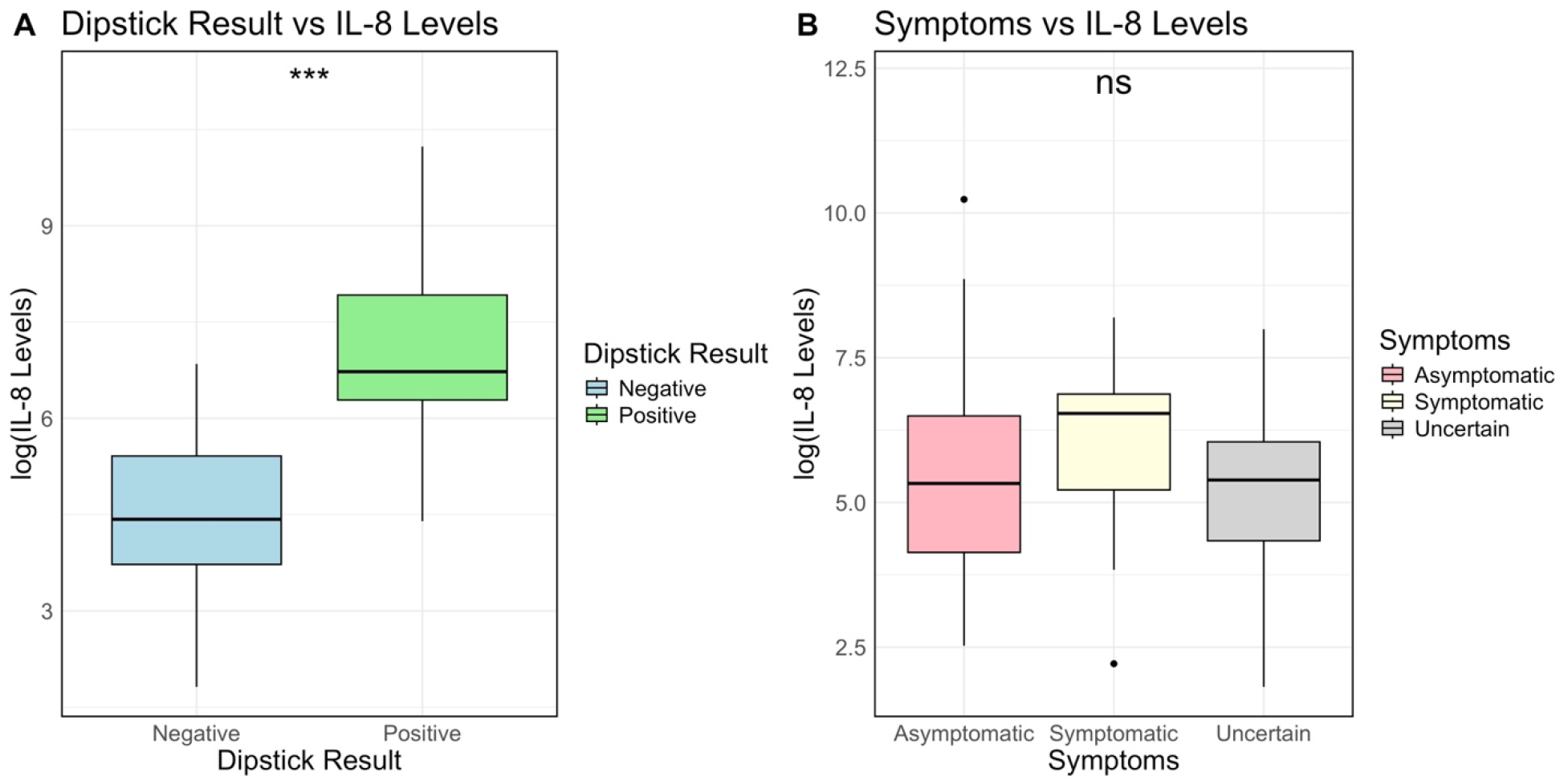
Association of urinary inflammatory marker IL-8 with dipstick result and symptoms. The figure shows that higher urinary IL-8 levels are associated with an increased likelihood of a dipstick-positive result (p < 0.001). In contrast, no clear difference in IL-8 levels is observed between participants who self-report as asymptomatic and those reporting symptoms (symptomatic p = 0.6; uncertain p = 0.3, with asymptomatic as the reference category).

These findings suggest that dipstick positivity and high colony counts of known uropathogens, particularly *Escherichia* and *Klebsiella*, are markers of biologically relevant inflammation in PLWD. Observed association with *Staphylococcus* and *Enterococcus* warrants caution, given these genera include both low-virulence skin and gastrointestinal commensals, respectively, and, less frequently, established urinary pathogens [26–28].

## Discussion

In this study, we characterised the nature and frequency of UTIs in PLWD using a longitudinal routine sampling method. This is the first study to do so in a cohort of PLWD living at home, a demographic less often studied due to the logistical challenges of working with dispersed populations outside care facilities.

We found UTIs to be more common in this group than in the general population, consistent with previous findings [29]. Notably, there was considerable variation in infection frequency: some individuals experienced persistent UTIs despite treatment, while others remained infection-free. This disparity may reflect differences in comorbidities such as diabetes, incontinence, or immunosuppressive therapy [30, 31]. The most frequently isolated bacteria—especially *Escherichia*—were consistent with known UTI pathogens [19].

Although diagnoses were primarily based on laboratory criteria, our analysis revealed that self-reported symptoms in PLWD showed poor correlation with both microbiological evidence (>10^5^ CFU/mL) and the presence of known uropathogens. In the general population, symptoms such as dysuria, urgency, and frequency are moderately predictive [32, 33], but cognitive impairments in PLWD likely impair symptom recognition and reporting [34]. This raises concerns that strict symptom-based guidelines, such as those from NICE, may lead to missed infections or inappropriate treatment of asymptomatic bacteriuria (ASB), contributing to antimicrobial resistance [19].

While dipstick testing has limited diagnostic reliability [35, 36], its rapidity and low cost make it a potentially useful initial screen when paired with confirmatory cultures. This contrasts with Public Health England’s recommendation against dipstick use in adults over 65 due to ASB prevalence [37]. However, our data suggest dipstick results in PLWD correlate more closely with both high bacterial loads and the inflammatory marker IL-8 than with reported symptoms. A two-step protocol, where a positive dipstick prompts culture testing, may help identify true infections while reducing unnecessary antibiotic use. This approach supports the need for dementia-specific diagnostic frameworks that balance sensitivity with the risk of overtreatment.

### Implications and Future Directions

Our findings underscore the need to refine UTI diagnostics for PLWD while cautioning against interventions lacking proven clinical benefit. Although dipsticks may help identify high bacterial loads, their impact on outcomes such as hospitalisation or delirium remains uncertain. Future studies should assess the effectiveness of dipstick-guided multi-step protocols in this regard.

Crucially, biomarker discovery is needed to differentiate true UTIs from ASB, as prior attempts to treat ASB based solely on bacterial persistence have not improved outcomes [18]. Biomarkers linked to host immune response (e.g IL-8 and C-reactive protein) offer more promise than bacterial presence alone. Although resource-intensive, such research could reduce diagnostic uncertainty and support better antibiotic stewardship.

## Data Availability

The data collected during the current study and the code used to analyse the data are available from a Zenodo public repository at (10.5281/zenodo.15720387).

https://doi.org/10.5281/zenodo.15720387

## Abbreviation

ASB: Asymptomatic Bacteriuria
CFU/mL: Colony-Forming Units per Millilitre
CI: Confidence Interval
CMHT-OP: Community Mental Health Teams for older adults
FDR: False Discovery Rate
GLMM: Generalized Linear Mixed-Effects Model
HFP PCN: Hammersmith & Fulham Partnership Primary Care Network
IL-8: Interleukin 8
IQR: Interquartile Range
LMM: Linear Mixed-Effects Model
SMMSE: Standardised Mini-Mental State Examination
NICE: National Institute for Health and Care Excellence
OR: Odds Ratio
PEA: Proximity Extension Assay
PLWD: People Living with Dementia
SABP: Surrey and Borders Partnership NHS Foundation Trust
UKDRI: UK Dementia Research Institute
UTI: Urinary Tract Infection

## Declarations

### Ethics approval and consent to participate

This study was submitted to, and approved by, the Surrey and Borders NHS Trust Research Ethics Committee (reference number: 19/LO/0102) and registered via the Integrated Research Application System (IRAS project ID: 257561). The research was conducted in accordance with the ethical principles set out in the Declaration of Helsinki. Informed consent was obtained from all participants prior to enrolment. Individuals unable to provide informed consent were excluded from participation.

#### Clinical Trial

Clinical trial number: not applicable.

### Consent for publication

Not applicable.

## Availability of data and materials

The data collected during the current study and the code used to analyse the data are available from a Zenodo public repository at (<u>10.5281/zenodo.15720387</u>).

## Competing interests

The authors declare that they have no competing interests.

## Funding sources

This work is supported by the UK Dementia Research Institute [award number UKI DRI-WBCN_PA5470], through UK DRI Ltd, principally funded by the Medical Research Council, and additional funding partners Alzheimer’s Society.

## Authors’ contributions

R.J. and R.C. are joint first authors, having both contributed equally to writing the main manuscript text, conducting the data analysis, and developing the R code. M.T., K.J., A.J.W., L.P.C., and M.A.C. provided laboratory support, including sample collection and processing for culture growth assays, dipstick tests, 16S Sanger sequencing, and Olink analyses. R.N. and D.W. provided clinical guidance and were responsible for diagnosing patients. D.J.W. and P.S.F. contributed by securing funding for the research. All authors reviewed the manuscript.

## Acknowledgements

We thank the patients and study partners who took part in the study. We also thank all members of the UK Dementia Research Institute Care Research & Technology Centre who contributed in some way to this work; a full list of members can be found in the supplementary file *“Centre Members”*. We are grateful to the Surrey and Borders Partnership, the sponsors of this study.

## Notes

### Competing Interest Statement

The authors have declared no competing interest.

### Funding Statement

This work is supported by the UK Dementia Research Institute which receives its funding from UK DRI Ltd funded by the UK Medical Research Council, Alzheimers Society, and Alzheimers Research UK.

### Author Declarations

The London-Surrey Borders Research Ethics Committee gave ethical approval for this work (REC: 19/LO/0102) The study is registered with National Institute for Health and Care Research (NIHR) in the United Kingdom under Integrated Research Application System (IRAS) registration number 257561.

### Summary of Updates

The authorship was expanded to include Rory Cave as joint first author. The abstract was refined, shifting from diagnostic accuracy to a broader and more comprehensive study of the epidemiology and diagnostic challenges of urinary tract infections (UTIs) in people living with dementia (PLWD). The methods section was updated to include greater detail on the experimental procedures, inclusion and exclusion criteria, statistical models, biomarker analysis, and an updated version of UTI classification. The description of molecular methods now specifies PCR conditions, sequencing workflows, and analytical pipelines, while the statistical framework incorporates generalised and linear mixed-effects models (GLMMs and LMMs) with false discovery rate (FDR) correction. In the results, the cohort description was refined from 91 to 86 participants following stricter inclusion criteria. Figures were renumbered and redrawn to improve visual clarity, with new analyses added, such as IL-8 biomarker quantification and taxon-specific modelling linking uropathogens to inflammation. Statistical correlations between dipstick results, bacterial load, and IL-8 concentration were also included. New subsections on microbial taxonomy, UTI frequency variability, and sex-related effects were introduced, enhancing the biological and clinical interpretation of findings. The discussion highlights the diagnostic inadequacy of symptom-based UTI detection in PLWD, supports re-evaluating dipstick use despite Public Health England guidance, and introduces IL-8 as a potential biomarker. New sections were added, including Ethics approval, data availability (Zenodo DOI), funding details, and comprehensive author contributions. The reference list was expanded and reformatted to align with academic journal standards.

## References

1. https://www.alzheimers.org.uk/sites/default/files/2018-05/Counting_the_cost_report.pdf. https://www.alzheimers.org.uk/sites/default/files/2018-05/Counting_the_cost_report.pdf. Accessed 23 Jun 2025.

2. National Mental Health D and NIN. Reasons why people with dementia are admitted to a general hospital in an emergency. 2015. https://webarchive.nationalarchives.gov.uk/ukgwa/20170302124526mp_/ http://www.yhpho.org.uk/default.aspx?RID=207311. Accessed 23 Jun 2025.

3. Counting_the_cost_report.pdf.

4. Corrado O, Swanson B, Hood C, Morris A, Ofili S, Capistrano J, et al. National Audit of Dementia Care in General Hospitals 2018–2019 Round Four Audit Report. https://www.rcpsych.ac.uk/docs/default-source/improving-care/ccqi/national-clinical-audits/national-audit-of-dementia/r4-resources/reports---core-audit/national-audit-of-dementia-round-4-report-online.pdf. Accessed 16 Apr 2025.

5. Phelan EA, Borson S, Grothaus L, Balch S, Larson EB. Association Between Incident Dementia and Risk of Hospitalization. Jama. 2012;307:165–72.

6. Gharbi M, Drysdale JH, Lishman H, Goudie R, Molokhia M, Johnson AP, et al. Antibiotic management of urinary tract infection in elderly patients in primary care and its association with bloodstream infections and all cause mortality: population based cohort study. BMJ. 2019;364:l525.

7. Eriksson I, Gustafson Y, Fagerström L, Olofsson B. Urinary tract infection in very old women is associated with delirium. Int Psychogeriatr. 2011;23:496–502.

8. Perry VH, Cunningham C, Holmes C. Systemic infections and inflammation affect chronic neurodegeneration. Nat Rev Immunol. 2007;7:161–7.

9. Dunn N, Mullee M, Perry VH, Holmes C. Association between Dementia and Infectious Disease: Evidence from a Case-Control Study. Alzheimer Dis Assoc Disord. 2005;19:91.

10. EAU Guidelines on Urological Infections - Uroweb. https://uroweb.org/guidelines/urological-infections. Accessed 16 Apr 2025.

11. Urinary tract infection: diagnostic tools for primary care. GOV.UK. 2024. https://www.gov.uk/government/publications/urinary-tract-infection-diagnosis. Accessed 16 Apr 2025.

12. Berman P, Hogan DB, Fox RA. The atypical presentation of infection in old age. Age Ageing. 1987;16:201–7.

13. Bai AD, Bonares MJ, Thrall S, Bell CM, Morris AM. Presence of urinary symptoms in bacteremic urinary tract infection: a retrospective cohort study of Escherichia coli bacteremia. BMC Infect Dis. 2020;20:781.

14. Cerejeira J, Lagarto L, Mukaetova-Ladinska EB. Behavioral and Psychological Symptoms of Dementia. Front Neurol. 2012;3:73.

15. Boscia JA, Kobasa WD, Knight RA, Abrutyn E, Levison ME, Kaye D. Epidemiology of bacteriuria in an elderly ambulatory population. Am J Med. 1986;80:208–14.

16. Nicolle LE. Asymptomatic bacteriuria in the elderly. Infect Dis Clin North Am. 1997;11:647–62.

17. Abrutyn E, Mossey J, Berlin JA, Boscia J, Levison M, Pitsakis P, et al. Does asymptomatic bacteriuria predict mortality and does antimicrobial treatment reduce mortality in elderly ambulatory women? Ann Intern Med. 1994;120:827–33.

18. Nicolle LE, Mayhew WJ, Bryan L. Prospective randomized comparison of therapy and no therapy for asymptomatic bacteriuria in institutionalized elderly women. Am J Med. 1987;83:27–33.

19. Diagnosis of urinary tract infections: quick reference tools for primary care. GOV.UK. https://www.gov.uk/government/consultations/urinary-tract-infection-diagnostic-tools-for-primary-care/diagnosis-of-urinary-tract-infections-quick-reference-tools-for-primary-care. Accessed 16 Apr 2025.

20. Roberts KB, Wald ER. The Diagnosis of UTI: Colony Count Criteria Revisited. Pediatrics. 2018;141:e20173239.

21. Barratt J. What to do with patients with abnormal dipstick urinalysis. Medicine (Baltimore). 2007;35:365–7.

22. Gu J, Chen X, Yang Z, Bai Y, Zhang X. Gender differences in the microbial spectrum and antibiotic sensitivity of uropathogens isolated from patients with urinary stones. J Clin Lab Anal. 2021;36:e24155.

23. Magliano E, Grazioli V, Deflorio L, Leuci AI, Mattina R, Romano P, et al. Gender and Age-Dependent Etiology of Community-Acquired Urinary Tract Infections. Sci World J. 2012;2012:349597.

24. Flores-Mireles AL, Walker JN, Caparon M, Hultgren SJ. Urinary tract infections: epidemiology, mechanisms of infection and treatment options. Nat Rev Microbiol. 2015;13:269–84.

25. Ko YC, Mukaida N, Ishiyama S, Tokue A, Kawai T, Matsushima K, et al. Elevated interleukin-8 levels in the urine of patients with urinary tract infections. Infect Immun. 1993;61:1307–14.

26. Becker K, Heilmann C, Peters G. Coagulase-negative staphylococci. Clin Microbiol Rev. 2014;27:870–926.

27. Ehlers S, Merrill SA. Staphylococcus saprophyticus Infection. In: StatPearls. Treasure Island (FL): StatPearls Publishing; 2025.

28. Codelia-Anjum A, Lerner LB, Elterman D, Zorn KC, Bhojani N, Chughtai B. Enterococcal Urinary Tract Infections: A Review of the Pathogenicity, Epidemiology, and Treatment. Antibiotics. 2023;12:778.

29. Rodriguez-Mañas L. Urinary tract infections in the elderly: a review of disease characteristics and current treatment options. Drugs Context. 2020;9:2020–4–13.

30. Geerlings SE. Urinary tract infections in patients with diabetes mellitus: epidemiology, pathogenesis and treatment. Int J Antimicrob Agents. 2008;31 Suppl 1:S54–57.

31. Tandogdu Z, Cai T, Koves B, Wagenlehner F, Bjerklund-Johansen TE. Urinary Tract Infections in Immunocompromised Patients with Diabetes, Chronic Kidney Disease, and Kidney Transplant. Eur Urol Focus. 2016;2:394–9.

32. Gágyor I, Rentzsch K, Strube-Plaschke S, Himmel W. Psychometric properties of a self-assessment questionnaire concerning symptoms and impairment in urinary tract infections: the UTI-SIQ-8. BMJ Open. 2021;11:e043328.

33. Giesen LG, Cousins G, Dimitrov BD, van de Laar FA, Fahey T. Predicting acute uncomplicated urinary tract infection in women: a systematic review of the diagnostic accuracy of symptoms and signs. BMC Fam Pract. 2010;11:78.

34. Hodgson N, Gitlin LN, Winter L, Czekanski K. Undiagnosed Illness and Neuropsychiatric Behaviors In Community-residing Older Adults with Dementia. Alzheimer Dis Assoc Disord. 2011;25:109–15.

35. Mambatta AK, Jayarajan J, Rashme VL, Harini S, Menon S, Kuppusamy J. Reliability of dipstick assay in predicting urinary tract infection. J Fam Med Prim Care. 2015;4:265–8.

36. Kristensen LH, Winther R, Colding-Jørgensen JT, Pottegård A, Nielsen H, Bodilsen J. Diagnostic accuracy of dipsticks for urinary tract infections in acutely hospitalised patients: a prospective population-based observational cohort study. BMJ Evid-Based Med. 2025;30:36–44.

37. Joseph A. The Diagnosis and Management of UTI in >65s: To Dipstick or Not? The Argument Against Dipsticks. Infect Prev Pract. 2020;2:100063.

