## Supplementary Figures for "A Cohort Study of Urinary Tract Infections in People Living with Dementia: Epidemiology and Diagnostic Challenges"

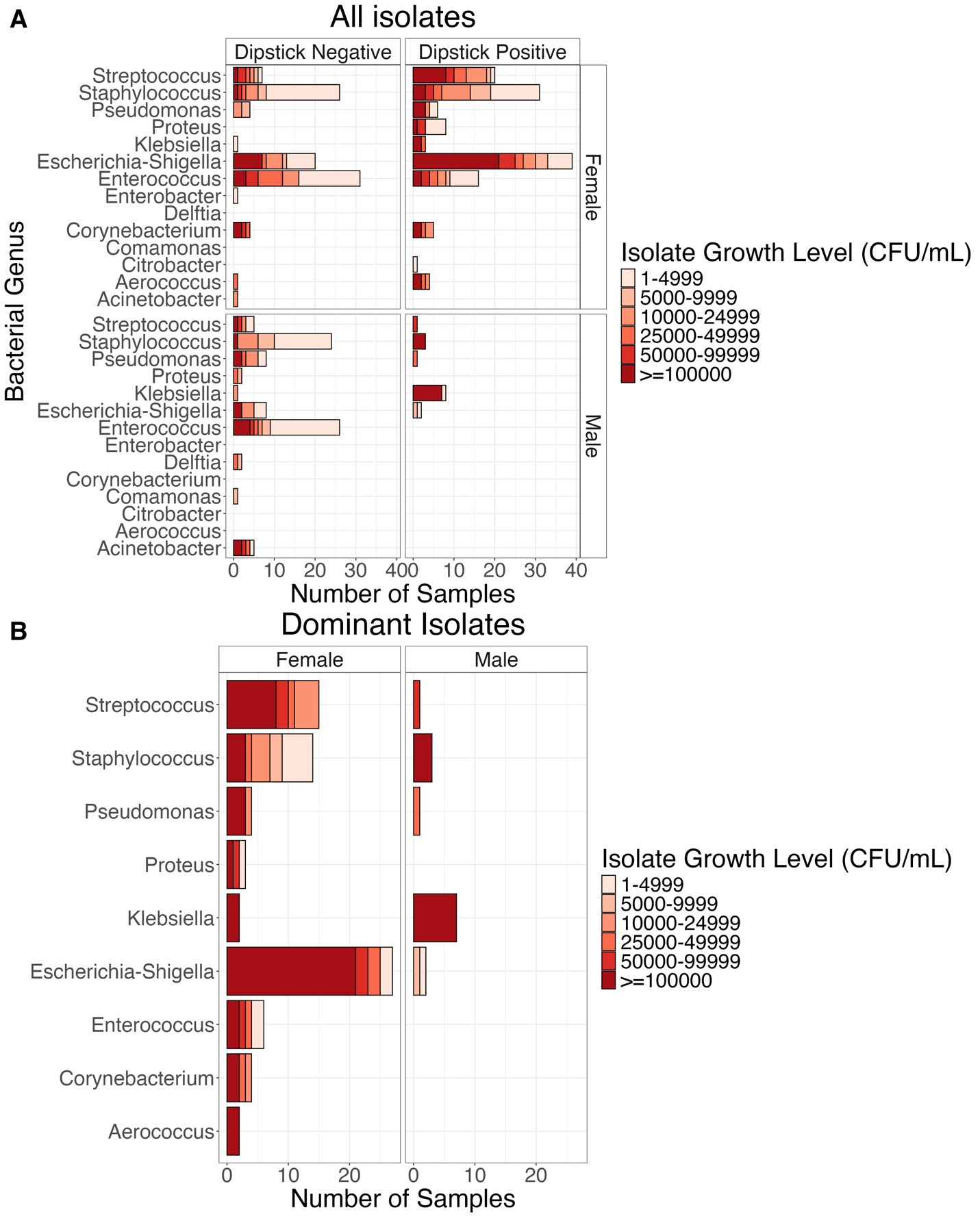

***Figure S1: Overview of Bacterial Taxa Identified in Urine Cultures****. (A) Bar plot illustrating the taxonomic identity and culture abundance of bacterial isolates from each chromogenic agar plate, stratified by sex and dipstick test result (leukocyte/nitrite positive).
(B) Bar plot highlighting the most abundant bacterial taxa isolated from leukocyte- and/or nitrite-positive samples, grouped by sex and coloured by culture abundance.
The figure shows that uropathogenic* Escherichia-Shigella *is among the most frequently identified taxa, often presenting with high colony counts exceeding 10⁵ CFU/mL (red tiles), and is more commonly observed in dipstick-positive samples compared to dipstick-negative ones.*

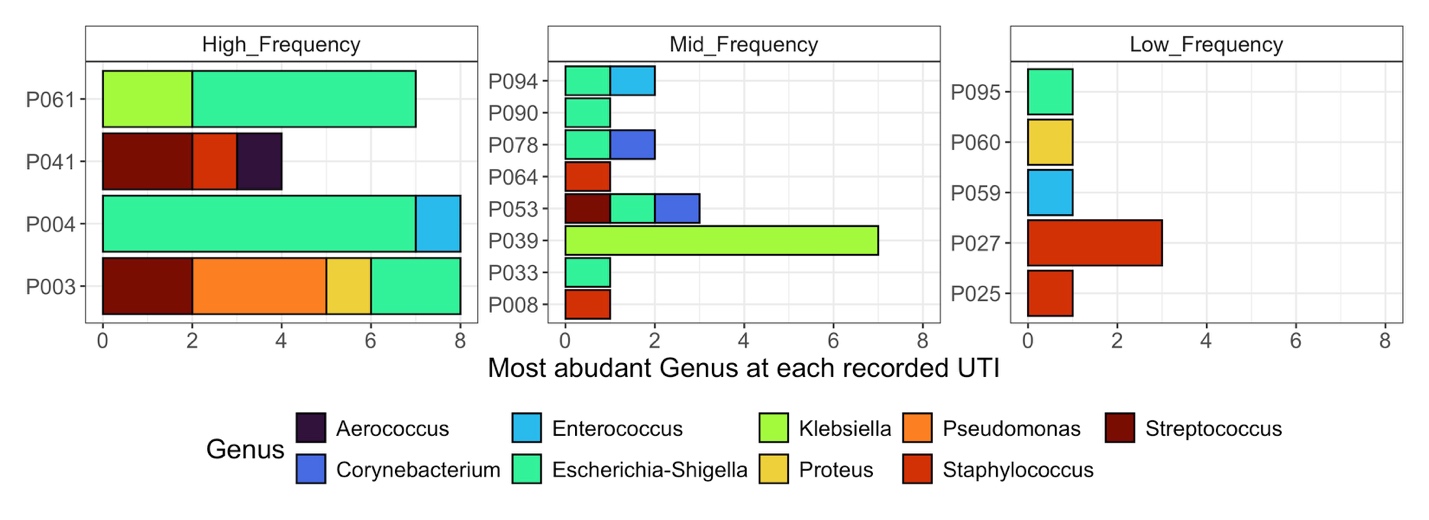

***Figure S2: Bacterial Taxa Associated with UTIs by Participant.*** *Bar plot displaying the taxonomic identity of bacterial isolates associated with urinary tract infection (UTI) in individual participants, grouped according to their overall UTI frequency. The figure demonstrates no clear difference in the bacterial taxa causing UTIs across participants with varying frequencies of infection.*
